# Towards integration of ^64^Cu-DOTA-Trasztusumab PET-CT and MRI with mathematical modeling to predict response to neoadjuvant therapy in HER2+ breast cancer

**DOI:** 10.1101/2020.06.10.20127639

**Authors:** Angela M. Jarrett, David A. Hormuth, Vikram Adhikarla, Prativa Sahoo, Daniel Abler, Lusine Tumyan, Danial Schmolze, Joanne Mortimer, Russell C. Rockne, Thomas E. Yankeelov

## Abstract

While targeted therapies exist for human epidermal growth factor receptor 2 positive (HER2+) breast cancer, HER2+ patients do not always respond to therapy. We present the results of utilizing a biophysical mathematical model to predict tumor response for two HER2+ breast cancer patients treated with the same therapeutic regimen but who achieved different treatment outcomes. Quantitative data from magnetic resonance imaging (MRI) and ^64^Cu-DOTA-Trastuzumab positron emission tomography (PET) are used to estimate tumor density, perfusion, and distribution of HER2-targeted antibodies for each individual patient. MRI and PET data are collected prior to therapy, and follow-up MRI scans are acquired at a midpoint in therapy. Given these data types, we align the data sets to a common image space to enable model calibration. Once the model is parameterized with these data, we forecast treatment response with and without HER2-targeted therapy. By incorporating targeted therapy into the model, the resulting predictions are able to distinguish between the two different patient responses, increasing the difference in tumor volume change between the two patients by >40%. This work provides a proof-of-concept strategy for processing and integrating PET and MRI modalities into a predictive, clinical-mathematical framework to provide patient-specific predictions of HER2+ treatment response.

## 1. Introduction

There is growing evidence that imaging-informed, mechanism-based mathematical models can accurately predict the development of cancers of the kidney [1], brain [2-5], lung [6, 7], and pancreas [8-11]. Importantly, these studies often aim to evaluate tumor growth or response to therapy on a patient-specific basis. A particular example of an imaging modality that can and has been utilized for mathematical modeling of tumor response is magnetic resonance imaging (MRI), which can be used to quantitatively characterize (for example) blood flow, vessel permeability, tissue volume fractions, cellularity, pH, and pO_2_ [12]. Additionally, positron emission tomography (PET) can quantitatively characterize molecular events related to (for example) metabolism, proliferation, hypoxia, and various cell surface receptors [13]. The strength of these imaging measurements is that they can be collected (with minimal invasion) at the time of diagnosis and then at multiple time points throughout treatment. Furthermore, imaging allows mathematical models to be initialized and constrained with patient-specific data rather than, for example, population data from the literature or animal data. Therefore, the ability to parameterize models with data that are readily accessible and specific to the individual enables mathematical modelling to potentially be integrated into clinical trials and, ultimately, translated to clinical practice.

For the particular case of breast cancer, we have previously developed mathematical models using patient-specific MRI data to calibrate the model’s parameters to the unique characteristics of each patient [14-20], thereby enabling patient-specific predictions. Recently, we presented a proof-of-principle study that extended a model to include estimates of drug delivery to each voxel *via* dynamic contrast-enhanced MRI (DCE-MRI), enabling a more accurate assessment of local tumor cell death due to therapy on a patient-specific basis [20]. Further assessment of this model’s predictive ability revealed that it could reliably distinguish between patients that would respond or not to therapy regimens—as defined by the response evaluation criteria in solid tumors (RECIST) [21]. However, the model did not perform as well for patients who also received targeted therapies in addition to chemotherapies. Specifically, for patients with tumors that overexpress the human epidermal growth factor receptor 2 (HER2) treated with antibodies targeted to this receptor (i.e., trastuzumab and/or pertuzumab), the model was not able to reliably distinguish tumors that would or would not respond to neoadjuvant therapy (NAT)—regimens that occur prior to surgery. This strongly suggests that the model must be amended to incorporate the effects of targeted therapies. Of course, extending a model results in a larger number of free parameters, which, in turn, requires more data to initialize and constrain the model. It is the overall goal of this study to develop a multi-modal imaging acquisition and analysis protocol that would enable patient-specific predictions of the response of HER2+ breast cancer to combination targeted and non-targeted therapies.

Breast cancers that overexpress the HER2 protein have a uniquely aggressive natural history and determine candidacy for HER2-directed therapies [22]. Trastuzumab is a humanized antibody that binds to the extracellular domain of HER2 and prevents intracellular signaling for proliferation. The addition of pertuzumab further inhibits downstream signaling by preventing the heterodimerization of HER2 [23]. The determination of HER2 status is made on a pre-treatment tumor biopsy specimen, which is a small sample of a larger tumor and may not be representative of the entire tumor and provides minimal information about heterogeneity of HER2 expression. Moreover, heterogeneity of HER2 expression and distribution of the HER2 antibody may play a role in HER2-directed treatment failure, when a patient does not achieve a complete response to therapy (unrelated to drug resistance). Several imaging modalities, including PET and MRI, can be used to assess tumor heterogeneity, density, perfusion, and therapy delivery as potential factors in determining response to HER2-directed therapy *in vivo*, in individual patients. Radiolabeled trastuzumab, ^64^Cu-DOTA-trastuzumab (^64^Cu-DT), has been used as PET imaging agent to characterize the delivery of HER2 targeted therapies [24]. We have previously utilized ^64^Cu-DT-PET to estimate the spatial distribution of trastuzumab in women with metastatic HER2 positive breast cancer. In this study, we use pretreatment MRI and ^64^Cu-DT PET-CT to predict response at surgery to the combination of cytotoxic chemotherapy with the humanized monoclonal antibodies, trastuzumab and pertuzumab in women with locally advanced HER2+ breast cancer (clinical trial NCT02827877).

Here, we first describe two patients from NCT02827877, one of whom had a complete response to therapy and another who did not. Second, we present the details of the image processing and analysis to yield data types that can be directly incorporated into the mathematical model. Third, we present the mathematical model and strategy for implementation. Fourth, we present the results of the predictions for the model with and without the HER2 therapy component to provide early evidence on the relative importance of these terms. Finally, we place the results in the context of the field, and discuss limitations of the study, and identify future avenues of investigations at the interface of multi-modality imaging and mathematical oncology.

## 2. Methods

### 2.1 Patient cohort

Patients with locally advanced breast cancer were considered eligible from clinical trials provided that they had biopsy confirmation of HER2 overexpression, ECOG (Eastern Cooperative Oncology Group) performance status 0-2, normal cardiac function, a primary tumor ≥ 2 cm or axillary lymph nodes ≥ 2 cm in diameter, and planned neoadjuvant chemotherapy with trastuzumab, pertuzumab, docetaxel and carboplatin. Participants could not have evidence of metastatic disease as determined by ^18^F-fluorodeoxyglucose (^18^F-FDG-) PET/CT and could not have received prior HER2 directed therapy. The City of Hope Institutional Review Board approved the study, and all patients provided written consent before participating (NCT02827877).

Prior to institution of chemotherapy, breast MRI, ^18^F-FDG- and ^64^Cu-DT-PET were performed. Patients received intravenous trastuzumab, pertuzumab, docetaxel and carboplatin every 3 weeks for 6 cycles in the absence of disease progression or unacceptable toxicity. Response status was determined by surgical pathology after the completion of NAT. See Figure 1 for details on the study design with respect to the NAT regimens. In particular, a pathological complete response (pCR) is defined as finding no viable tumor cells present in the primary tumor or lymph nodes at the time of surgery following completion of neoadjuvant systemic therapy.

**Figure 1.**
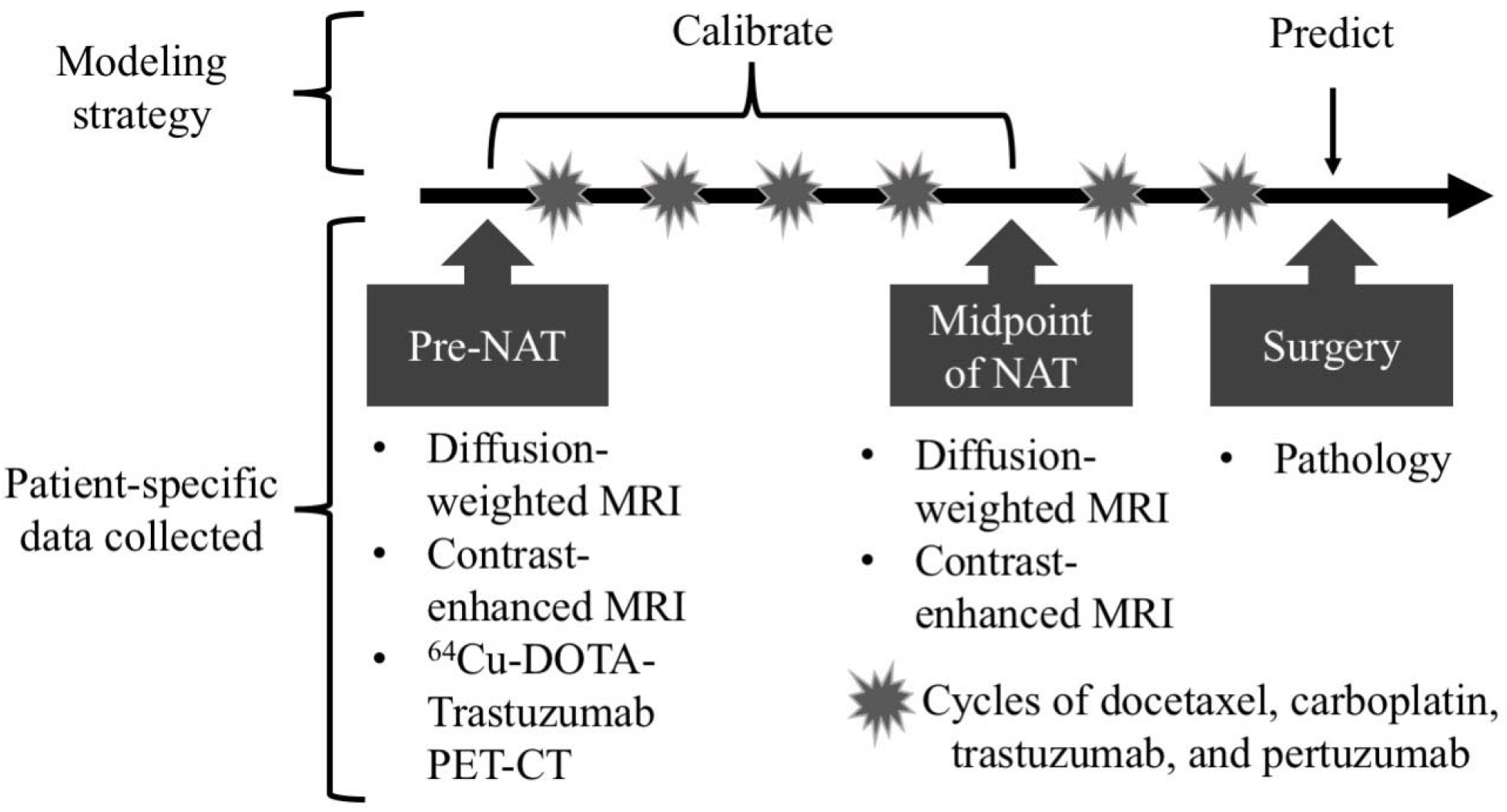
Schematic of the integrated mathematical-experimental approach employed in the study. Prior to NAT, several imaging and biopsy data types were collected for each individual patient, including ^18^F-FDG PET-CT, but it is not being utilized in this current study. At a midpoint of NAT, a follow-up MRI scans were also performed. At surgery, tissue was collected and sent to pathology for evaluation. The first two sets of data (pre-NAT and at the midpoint of NAT) were used to calibrate the mathematical model’s parameters for each individual patient. With the patient-specific parameters, the model was reinitialized at each patient’s second imaging session data and run forward to predict tumor status at the time of surgery. Then the model’s predictions were compared to the clinical outcome determined at the time of surgery.

Prior to therapy, a diagnostic biopsy was performed. For both patients, their diagnostic biopsies received an immunohistochemistry (IHC) HER2-overexpression score of 3+ (scores range from 0 to 3+ for different intensity levels of IHC staining), indicating that the tumors display complete, intense circumferential membranous staining in > 10% of tumor cells. Further inspection and processing of the biopsy slides supports that the cancer cells have fairly uniform HER2 expression (see Supplemental Materials for images of the IHC staining results for each patient), an important point we return to in the discussion.

In this study, to illustrate our multi-modal imaging based mathematical modeling approach, we selected two patients from NCT02827877 with contrasting tumor responses; where, at the time of surgery, one patient achieved a pCR (patient 1) while the second patient had residual disease (non-pCR, patient 2).

### 2.2 MRI data acquisition

MRI data were acquired at baseline after diagnosis but prior to treatment and also at a midpoint of NAT prior to surgery (for patient 1, after four cycles of therapy and for patient 2, after three cycles of therapy). MRI was performed using a 1.5T GE scanner with the body coil and a four-channel bilateral phased-array breast coil as the transmitter and receiver, respectively. Each MRI session began with pilot scans, axial *T*_*2*_-weighted MRI with fat-saturation, and axial *T*_*1*_-weighted without fat-saturation, before proceeding to diffusion-weighted (DW)-MRI and contrast enhanced (CE)-MRI and sagittal delayed *T*_1_ weighted acquisitions. The bilateral DW-MRI was acquired in the axial plane with a single-shot spin echo planar imaging sequence, with *b*-values of 0, 600, 800, 1000 and 1500 s/mm^2^, TR/TE = 6675 ms/97.3 ms, and 5 signal averages. The acquisition matrix was 128 × 128 (reconstructed to 256 × 256) over a 360 × 360 mm^2^ field of view, and a slice thickness of 5 mm. Then bilateral, full breast, axial CE-MRI was acquired with a fat suppressed, gradient echo-based 3D VIBRANT (Volume Image Breast Assessment) sequence. CE-MRI acquisition parameters were TR/TE/α = 4 6.6 ms/3.19 ms/10°, and an acquisition matrix of 420 × 420 (reconstructed to 512 × 512) over a 340 × 340 mm^2^ field of view, and a slice thickness of 1.8 mm. The intravenous injection of Gd-BOPTA (Multihance, Bracco, Italy) was 0.2 mmol/kg at 2 ml/s delivered by a programmable power injector followed by a 20 ml saline flush. One baseline and three post contrast image volumes were acquired with temporal resolution of 180 sec per acquisition. The acquisition time for the CE-MRI was approximately 20 min, depending on the number of slices needed (112–120) in each image volume for full breast coverage. See Figure 2 for examples of both patient’s DW- and CE-MRI data.

**Figure 2.**
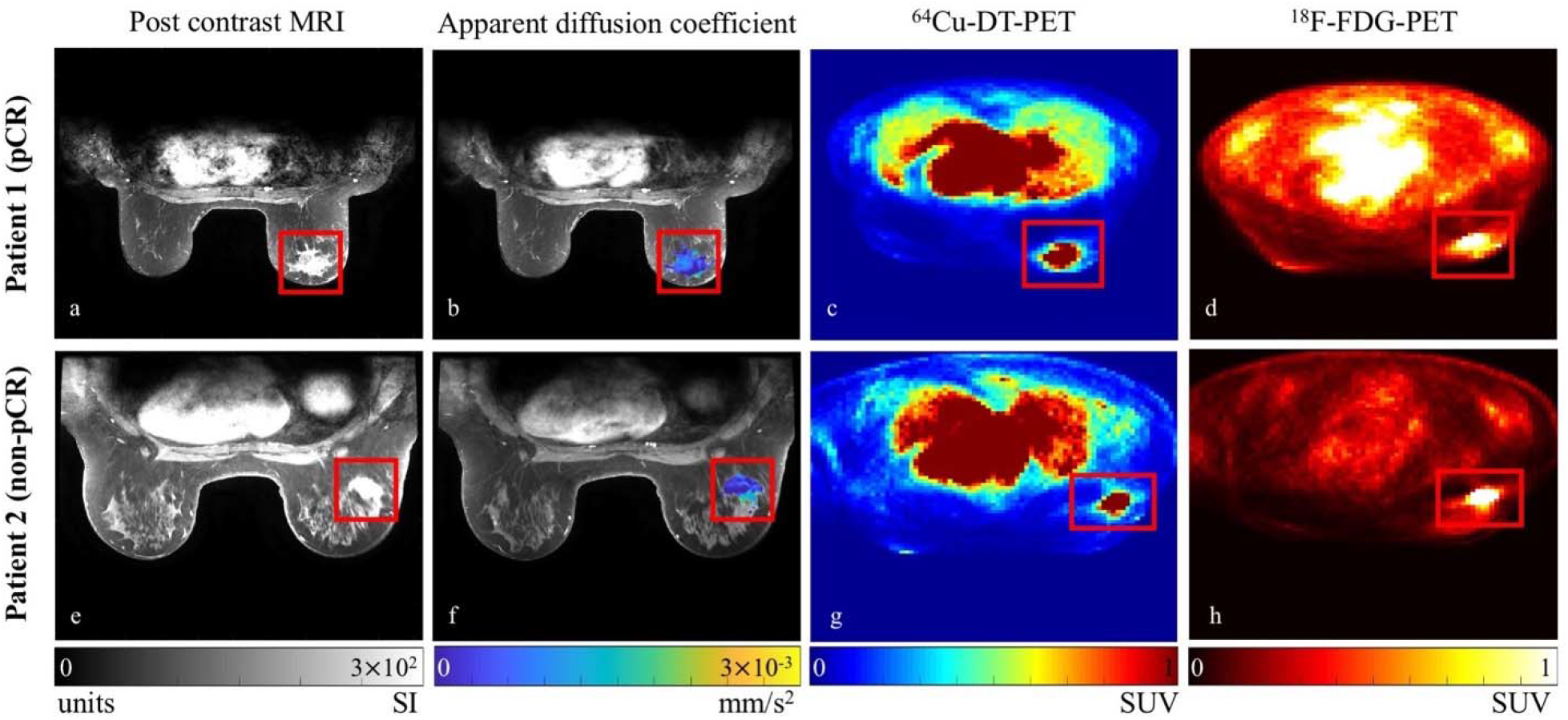
Example MRI and PET data for a central slice from both patients with the tumor region boxed in red. Note that MRI and PET data were acquired in the prone position as opposed to the usual supine position using our novel breast support device (see text for details), and these example images are prior to intra-scan registration that aligns the data to one common space for modeling. Note that in the CE-MRI data (panels (a) and (e)), the tumor enhances more than the surrounding tissues. The DW-MRI data is represented here by the calculated *ADC* map (overlain on an anatomical image) in panels (b) and (f). Note that low *ADC* values (blue) are indictive of areas of higher cellularity. Panels (c) and (g) show the ^64^Cu-DT-PET data, while panels (d) and (h) present the ^18^F-FDG-PET data. Note that due to the tumor location and arm positioning, for patient 2, part of her arm is out of view in the cropped image. For both a high signal intensity was observed within the tumors for the ^64^Cu-DT-PET data, whereas for the ^18^F-FDG-PET data the signal was not as strong for patient 1 for this slice. (While the ^18^F-FDG-PET data was collected with the data in this study and is part of the image processing pipeline, it was not used for this modeling study.)

### 2.3 PET data acquisition

PET data acquisition occurred at baseline after diagnosis but prior to treatment. Images were collected using a Discovery STE clinical PET/CT scanner (GE Healthcare, Waukesha, WI, USA). Data were acquired in the prone position using a custom-built padded support [25-28]. A low-mAs CT scan was acquired using the smart mA setting for attenuation correction of the emission data. Standard-of-care ^18^F-FDG PET/CT scan was acquired to evaluate patient eligibility.

^64^Cu was provided by the Mallinckrodt Institute of Radiology, Washington University School of Medicine). ^64^Cu radiolabeled trastuzumab was prepared according to an investigational new Drug Application (IND 109971). The procedure includes heating at 43^°^C for 45 minutes followed by incubation with an excess of diethylenetriaminepentaacetic acid, which eliminates ^64^Cu binding to secondary chelating sites on the antibody while maintaining the immunoreactivity of the radiolabeled product. The ^64^Cu-DT (trastuzumab dose, 5 mg) was mixed with saline (25 mL). Fifteen minutes prior to administration of the radiolabeled trastuzumab, 45 mg of trastuzumab was administered IV to decrease the hepatic uptake of ^64^Cu without affecting tumor uptake [25]. HER2 is expressed in both healthy and cancerous cells albeit to different extents; therefore, the administration of a cold dose of trastuzumab prior to imaging is required to saturate normal tissues taking up the antibody [29]. After 60-90 minutes post-injection of the cold antibody, patients receive 15 mCi of ^64^Cu-DT administered intravenously over 10 min and undergo a PET/CT scan within 28 - 30 hours. Attenuation and scatter corrected ^64^Cu-DT scans were reconstructed on a 128 × 128 matrix over a 700 mm square field of view using an ordered subset expectation maximization algorithm (GE VUE point HD) with two iterations and 20 subsets. Voxel resolution of the reconstructed image was 5.5 × 5.5 × 3.3 mm^3^. See Figure 2 for central slice examples of both patient’s ^64^Cu-DT- and ^18^F-FDG-PET data. While the standard-of-care ^18^F-FDG-PET data was collected, it was not used for this modeling study.

### 2.4 Image analysis

Our approach to imaging-based modeling requires that all image sets be registered to the same imaging space. Thus, the first step in the data processing protocol consists of registering the DW-MRI and CE-MRI data for each scan session for motion correction followed by registering the PET-CT data to the MRI data. For each patient, the MR images acquired at each session (intra-visit) were registered using a rigid algorithm where the diffusion-weighted images were linearly interpolated in 3D to match the resolution of the CE-MRI data and registered to the contrast enhanced images. The interpolation of images was implemented using MATLAB’s (MathWorks, Natick, MA) function *interp3* and the rigid algorithm used for the intra-visit registration *via* the function *imregister*. A deformable intra-visit registration was employed to align the PET/CT and MRI data *via* the MATLAB function *imregdemons*, where the field of view for the corresponding CT scan was trimmed and registered to the contrast enhanced images, and the resulting deformation field was applied to the PET images (linearly interpolated in 3D to match the CE-MRI resolution). See Figure 3 for example images of the ^64^Cu-DT-PET to MRI registration for patient 1.

**Figure 3.**
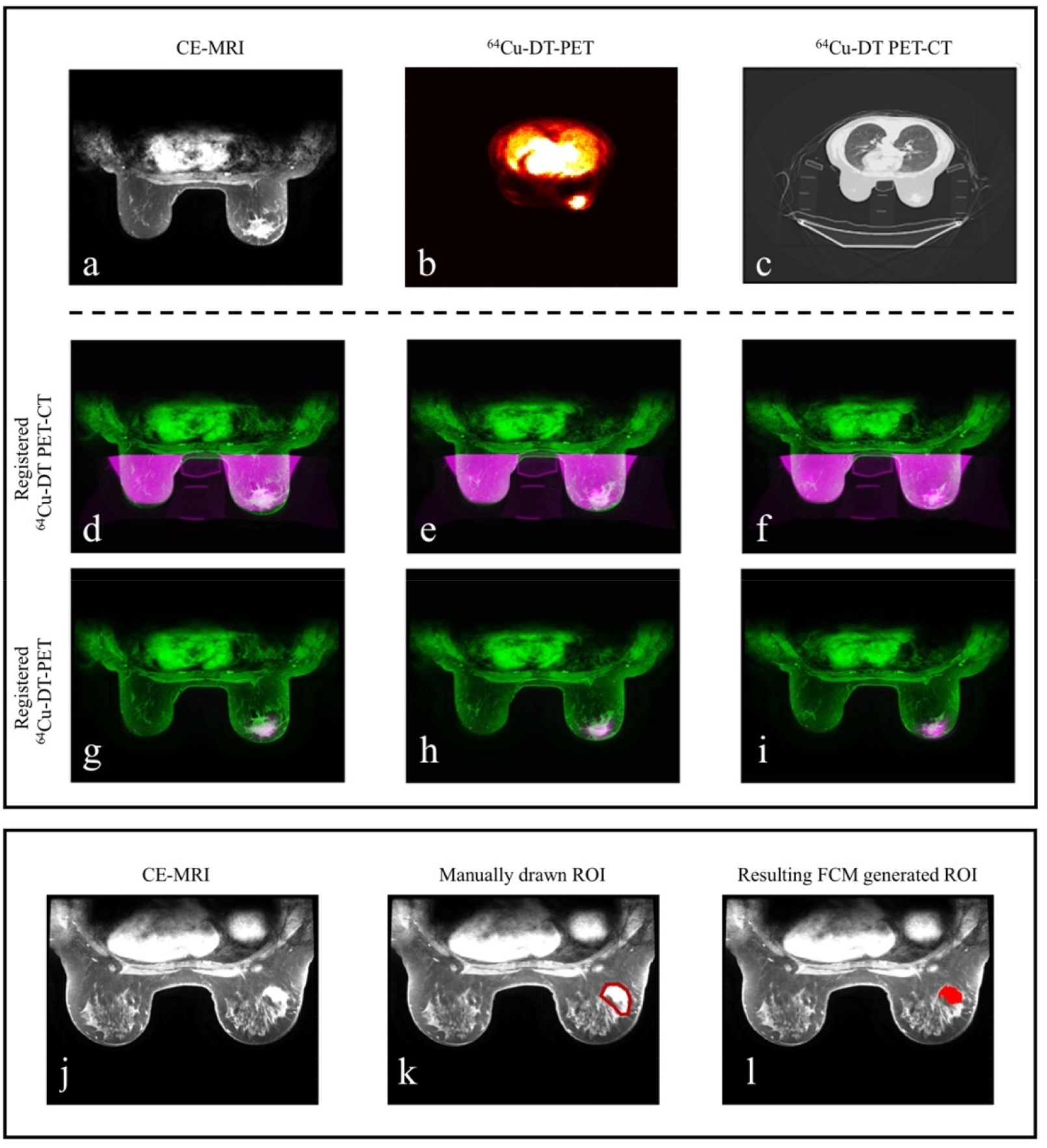
Example images of data processing for intra-scan registration of PET data to MRI data (upper panel) and generation of tumor ROIs (lower panel). Upper panel, deformable intra-scan registration for PET data to MRI data. Note that these images were all acquired in the prone position as opposed to the usual supine position using our novel breast support device (see text for details). The top row depicts a central slice for patient 1 at baseline for the CE-MRI (panel a), ^64^Cu-DT-PET (panel b), and the ^64^Cu-DT PET-CT (panel c). After the CT data is trimmed to only include breast tissue (i.e., the chest cavity and breast support was removed from the original images), the CT images were registered to the CE-MRI data using a fully deformable registration algorithm. Panels (d-i) depict the resulting overlap between the CE-MRI data and registered CT-PET data for three central slices (green represents the CE-MRI data and pink represents the cropped ^64^Cu-DT PET-CT and ^64^Cu-DT-PET for panels (d-f) and (g-i), respectively). Lower panel, example images for generating tumor ROIs from CE-MRI data using the FCM method (patient 2). Panel (j) depicts a central slice of the CE-MRI data. Panel (k) shows a manually drawn, conservative ROI (red) on the CE-MRI data. Panel (l) depicts the FCM generated ROI (red).

The second step encompasses the preliminary processing of the data including calculating the apparent diffusion coefficient (*ADC*) of water maps, identifying the tumor regions of interest (ROIs), and approximating the drug distributions for each patient. The *ADC* of water was calculated from the DW-MRI data using all five b-values *via* standard methods [31]. Using the CE-MRI data, a fuzzy c-means (FCM)-based algorithm is applied to a manually drawn, conservative ROI to identify the boundaries of the tumor [32]. The FCM algorithm is a clustering method that not only partitions the voxels into classes but also assigns a weighting based on probability of a voxel belonging to the tumor. See Figure 3 for an example of applying this approach to patient 2. Also using the CE-MRI data, to approximate the distribution of chemotherapy drug throughout the tissue for each patient, a normalized map of the vasculature was calculated by: (1^st^) subtracting the average baseline signal from the pre-contrast dynamics, (2^nd^) computing the area under the curve (AUC) for each voxel post injection of the contrast agent, and (3^rd^) dividing by the maximum AUC over the whole ROI. This normalized AUC map was used to define the initial systemic drug distribution throughout the tumor and surrounding tissue at the time of each therapy dose. The ^64^Cu-DT-PET data is used to define a normalized map of the distribution of the targeted anti-HER2 therapy trastuzumab based on the dosage of the radiotracer the patients received. See Figure 4 for the resulting drug distribution maps in the breast domain for a central slice of each patient’s tumor at baseline.

**Figure 4.**
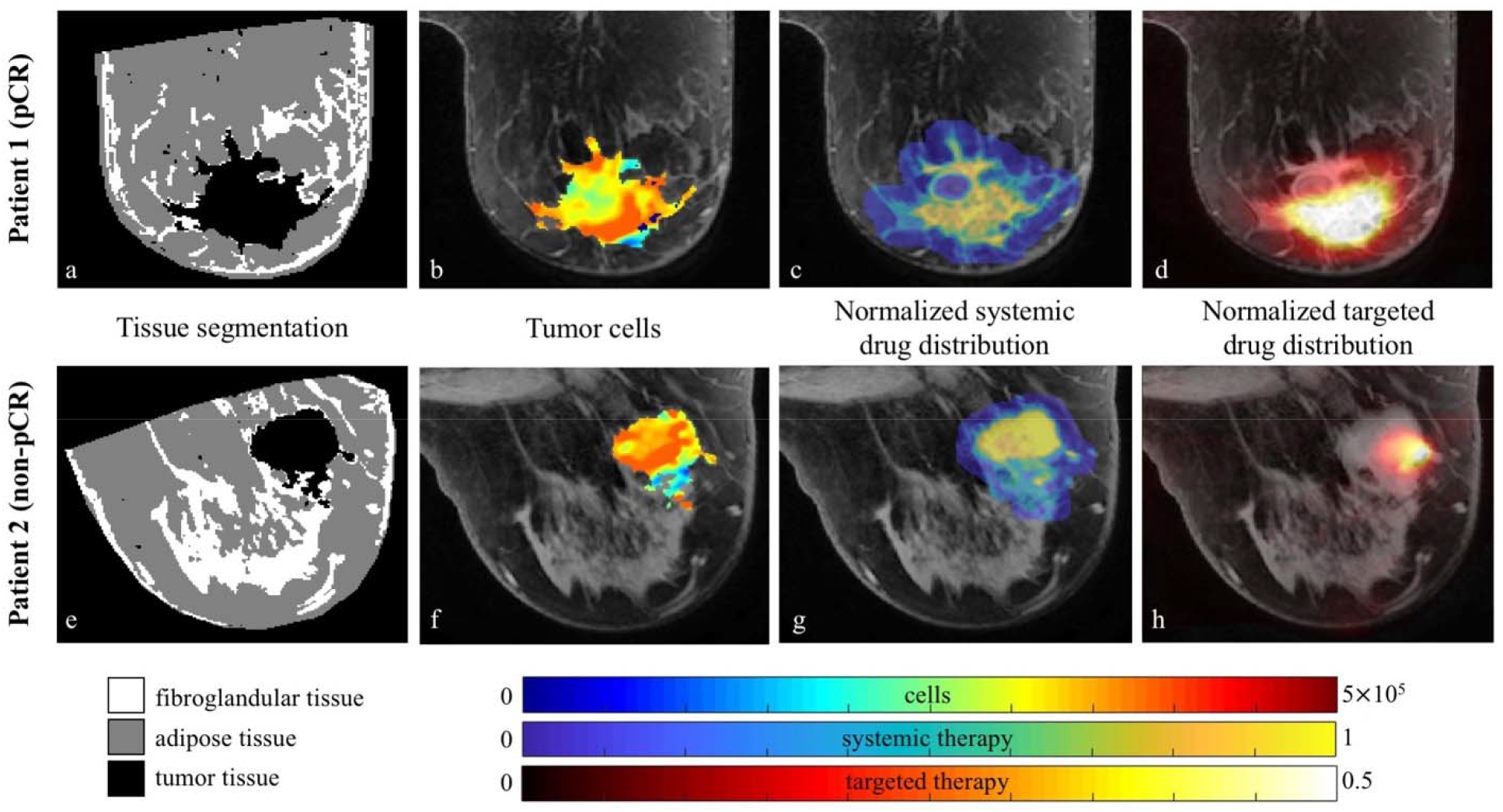
The central slide of each tumor for both patients are shown to depict the tissue segmentation (panels a and e), number of tumor cells (b and f), and the normalized estimates of the systemic (c and g) and targeted (d and h) therapies used in the model. The tissue segmentation identifies the tumor (dark area), fibroglandular (white), and adipose (grey) tissues within the breast. For the remaining columns, the parameter maps (i.e., the colored pixels) are overlaid on anatomical images of the breasts (grey). At surgery, patient 1 was designated as a pCR, while patient 2 had residual disease and was designated as non-pCR. Both patient’s tumor exhibit areas of high cellularity, and the approximate drug distribution of the systemic therapies have similar intensities. Comparing the targeted distribution maps, patient 1 appears to have had greater drug distribution compared to patient 2 for this central slice.

The third step is a registration that aligns the images and calculated maps of both of the patients’ scans (across time, inter-visit) to one common spatial coordinate system (co-registered). A non-rigid registration algorithm (an adaptive basis algorithm) with a constraint that preserves the tumor volumes at each time point was used [33]. This was accomplished using the open source toolbox Elastix [34, 35].

The final step is calculating the modeling quantities, including defining maps of the different tissues of the breast and calculating the number of tumor cells in the ROIs from the *ADC* of each voxel. The CE-MRI data were used for the tissue segmentation. The fibroglandular and adipose tissues were segmented using a k-means clustering algorithm that partitions the tissues of the breast into separate clusters. The resulting masks are used for the assignment of tissue stiffness properties in the mathematical model (see below). The *ADC* value for each voxel within the tumor (as segmented using the above methods) was converted to an estimate for the number of tumor cells per voxel at each 3D position 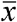 and time *t*, 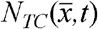, *via* our established methods [16-18, 20, 31]:

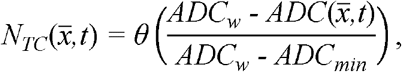

where *ADC*_*w*_ is the *ADC* of free water (3 × 10^−3^mm^2^/s at 37^°^ C) [36], 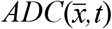 is the *ADC* value for the voxel at position *x* and time *t*, and *ADC*_*min*_ is the minimum *ADC* value over all tumor voxels for the patient [14, 37]. The parameter *θ* is the carrying capacity describing the maximum number of tumor cells that can physically fit within a voxel; its numerical value is determined by assuming a spherical packing fraction of 0.74 [38], a nominal tumor cell radius of 10 μm, and the voxel volume (2.18 mm^3^). See Figure 4 for the resulting tissue segmentation and tumor cell maps in the breast domain for a central slice of each patient’s tumor at baseline.

The tumor volume was approximated as the product of the total number of voxels within the segmented tumor ROI and the voxel volume. This measure of volume was also applied to all the modeling prediction results for direct comparison to the data. Note that for patient 1, there is no associated DW-MRI data for scan 2. We have confidence about the volume at the time point, so a tumor cell map was generated for the ROI at scan 2 to have a similar percent reduction in total tumor cellularity as achieved by patient 2.

### 2.5 Mathematical model

We have previously developed a 3D mathematical model that includes the mechanical coupling of tissue properties to tumor growth and the delivery of systemic therapy [20, 21]. This model was designed to be initialized with patient-specific, imaging data to the response of breast cancer patients to NAT [17-19]. The governing equation (a reaction-diffusion type partial differential equation) for the spatiotemporal evolution of tumor cells, 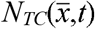, with respect to time, *t*, and per voxel, 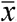, is:

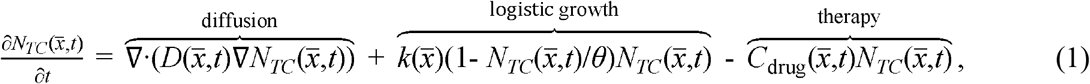

where the first term on the right-hand side describes the effects of tumor cell movement, the second term describes the growth of the cells, and the third term describes the effect of chemotherapy. All model parameters and functions are described in Table 1, and the reader is encouraged to refer to it as they move through the description of the mathematical model. This model has been well documented through its evolution [14-20], but we describe the established features (diffusion and growth) and discuss the expanded therapy term, 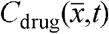, all together here. The first term on the right-hand side of Eq. (1), representing the random diffusion (movement) of the tumor cells, 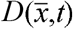, is mechanically linked to the breast tissue’s material properties *via*:

**Table 1.** Description of the variables and parameters for the model system including the assigned parameter values and specification of units.

| Variable | Description |
| --- | --- |
| $N_{TC}(\bar{x}, t)$ | Number of tumor cells in the voxel at position $\bar{x}$ at time $t$ |
| $D(\bar{x}, t)$ | Diffusion coefficient of tumor cells, where $D(\bar{x}, t) = D_0 \exp(-\gamma \sigma_{vm}(\bar{x}, t))$ (mm<sup>2</sup>/day) |
| $\sigma_{vm}$ | Von Mises stress (kPa) |
| $\vec{u}$ | Displacement vector due to tumor cell growth (mm) |
| $G$ | Shear modulus due to breast tissue properties, where $G = E/(2(1-\nu))$ (kPa) |
| $C_{tissue}^{drug}(\bar{x}, t)$ | Concentration of drug in the tissue in the voxel at position $\bar{x}$ at time $t$ (dimensionless) |

| Parameter | Description | Value |
| --- | --- | --- |
| $D_0$ | Diffusion coefficient of tumor cells without stress | Calibrated, mm<sup>2</sup>/day |
| $\gamma$ | Mechanical coupling coefficient for stress | Assigned at $2.0 \times 10^{-3}$ (1/kPa) |
| $\nu$ | Poisson's ratio | Assigned, 0.45 (dimensionless) |
| $E$ | Young's modulus for adipose, fibroglandular, and tumor tissues | Assigned, 4 kPa, 2kPa, and 20 kPa, respectively |
| $\lambda$ | Coupling constant for displacement of tumor cells | Assigned as $2.5 \times 10^{-3}$ (dimensionless) [39] |
| $k(\bar{x})$ | Proliferation rate of tumor cells per voxel | Calibrated, 1/day |
| $\theta$ | Carrying capacity of tumor cells in the voxel at position $\bar{x}$ | Calculated, $2.02 \times 10^6$ cells |
| $\alpha$ | Efficacy of the chemotherapy | Calibrated, 1/day |
| $\beta$ | Drug exponential decay rate | Calibrated, 1/day |
| $\mu$ | Efficacy of the targeted therapy | Calibrated, 1/day |

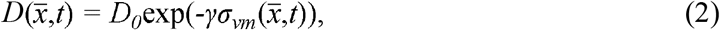

where *D*_0_ is the diffusion coefficient in the absence of external forces, and the exponential term damps *D*_*0*_ though the von Mises stress, 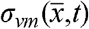, which is calculated for the fibroglandular and adipose tissues within the breast—where fibroglandular tissue has greater stiffness compared to adipose [39]. This mechanical coupling to the diffusion is subject to an equilibrium dependent upon changes in tumor cell number:

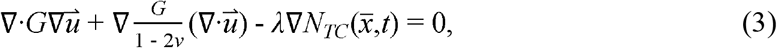

where *G* the shear modulus, where *G* = *E*/(2(1 - *ν*)) for the Young’s modulus (E) and Poisson’s ratio (*ν*) material properties, 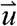 is the displacement due to tumor cell growth, and *λ* is another empirical coupling constant [17-19, 40-45]. Therefore, the diffusion term encompasses tumor changes such as growth or response to therapy that can cause deformations in the surrounding healthy tissues (i.e., fibroglandular and adipose tissues), thereby changing the stress field and the associated expansion of the tumor.

The second term on the right-hand side of Eq. (1) is the reaction term that describes tumor proliferation through logistic growth at the rate *k* and up to a carrying capacity, *θ*. The carrying capacity is defined per voxel using approximate cell size and packing density (as described above in the calculation of tumor cells, section 2.4), while the proliferation rate is calibrated per voxel for each individual patient (the numerical details on model calibration are detailed below in section 2.6).

The final term on the right-hand side of Eq. (1)—the therapy term—describes the spatiotemporal distribution of each systemic drug in the tissue and its effect on the cells of each voxel using the following equation,

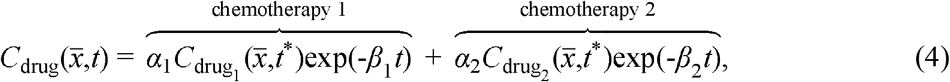

where *α*_i_ is the efficacy of each chemotherapy on the tumor cells; 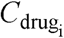 is the initial distribution of each drug for each dose (changing with time *t*_*_, described below), and the exponential decay terms, exp(-*β*_*i*_*t*), represent the eventual washout of drug over time after each dose. The *α*_i_ and *β*_*i*_ parameters are calibrated for each patient and chemotherapy, where the β_i_ calibration is restricted using bounds defined from ranges found in the literature for the terminal elimination half-lives of each drug [46-51]. Previously,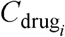, was approximated using a pharmacokinetic analysis of dynamic CE-MRI [20]; however, the present data set does not have the requisite temporal resolution or number of time points for such an analysis. Thus, we use the normalized AUC map (described in section 2.4) for each voxel post injection of the contrast agent. This drug distribution is dependent on the time *t* _*_, indicating that for the calibration of the model the drug distribution map is derived from scan 1, but an updated drug distribution map from scan 2 is provided to the model to predict the tumor at the time of surgery. Therefore, the drug effect in the tumor tissue is spatially non-uniform and temporally varying based on the individual patient’s response to therapy and NAT schedule.

Eqs. (1) – (4) do not account for targeted therapies. To overcome this limitation, Eq. (1) was modified to account for the change in proliferation rate due to trastuzumab and pertuzumab binding. As described in the introduction, the primary mechanism of action for these two targeted antibodies is the reduction in proliferation by binding to HER2, interrupting intracellular signaling. Therefore, similar to our previous *in vitro* modeling of trastuzumab [52], Eq. (1) becomes:

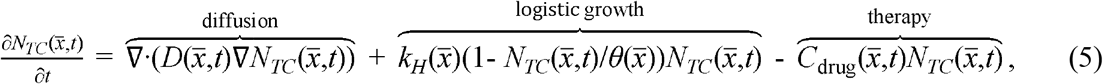

where *k*_*H*_ depends on the concentration of the targeted therapies,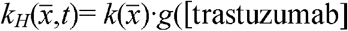. The proliferation *k*_*H*_ includes the spatially defined proliferation map 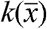 as well as a function dependent on the distribution of the two targeted drugs *g*(trastuzumab], [pertuzumab]). As the ^64^Cu-DT-PET data is specific to the trastuzumab antibody, we make the simplifying assumption that this data can be used to also approximate the distribution of pertuzumab and define the growth modulation function as:

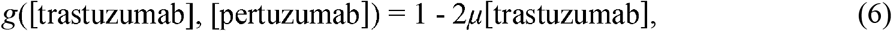

where [trastuzumab] and [pertuzumab] indicate the concentrations of trastuzumab and pertuzumab, respectively, and *μ* represents the effectiveness of the targeted therapies. Note that we assume that the targeted therapies have a constant effect once administered (i.e., drug clearance is not accounted for in this initial study, and we return to this important point in the Discussion section. Additionally, we assume the reduction in proliferation is linear with concentration and that the entire tumor burden may be affected by the targeted therapies. From here we refer to the model without the effect of the targeted therapy explicitly incorporated as the “MRI-based model” (i.e., Eqs. (1) – (4)), while the model that explicitly incorporates the targeted therapy as the “PET/MRI-based model” (i.e., Eqs. (2) – (6)). Note that the MRI-based model is the same as the PET/MRI-based model when the concentrations of the targeted therapies are set to zero.

### 2.6 Model parametrization and evaluation of predictions

All simulation codes and numerical calculations were written and executed in MATLAB (MathWorks, Natick, MA). The model was implemented in three dimensions (3D) with a fully explicit finite difference scheme with Δt = 0.25 day with the mesh dimensions defined by the size of the CE-MRI voxels. The size of the computational domain is set by a rectangle whose dimensions are determined by the size of the breast for each patient. A no flux boundary condition was prescribed at the boundary of the breast.

Our modeling approach used the two MRI data sets for each patient to calibrate the mathematical model and then simulate the model to the time of surgery to make a prediction of tumor response. See Figure 1 for a graphical depiction of the implementation of our modeling approach. Specifically, we used the cellularity maps of each tumor (derived from the *ADC* of the DW-MRI data—see section 2.4) to calibrate model parameters (*D*_*0*,_ *α, β*, and *µ* parameters are global; *k* is spatially determined). For the calibration, a Levenberg–Marquardt least squares non-linear optimization was used, where the sum of squared errors between the simulated tumor cell numbers from the model and the calculated number of tumor cells from the imaging data was minimized. To reduce computation time for all calibrations, the voxel matrix within this designated rectangular domain was down sampled by a factor of four. See [42] for additional details on the full development of these numerical methods.

Using these patient-specific, calibrated parameters, the model was reinitialized with the tumor cellularity, tissue, and drug distribution maps from scan 2 and run forward to the time of surgery to predict tumor response. We applied this approach with and without HER2-directed targeted therapy incorporated into the model, where the predictions of each model were evaluated by comparing two measures quantifying tumor response—total tumor cellularity and total tumor volume. Using these measures, the predicted percent change in the tumor responses from baseline (scan 1) to surgery as well as scan 2 to surgery were compared between the two models.

## 3 Results

Figure 5 depicts a central tumor slice of the resulting prediction for the time of surgery for each version of the model for both patients. Tumor cell number is overlaid in color on an anatomical image of the breast tissue. Notice that the original MRI-based model predicts larger tumors with higher cellularity for both patients compared to the PET/MRI-based model with the targeted therapy. In particular, for patient 1, the PET/MRI-based model predicts a tumor approximately 25% of the total tumor cellularity of that of the MRI-based model’s prediction.

**Figure 5.**
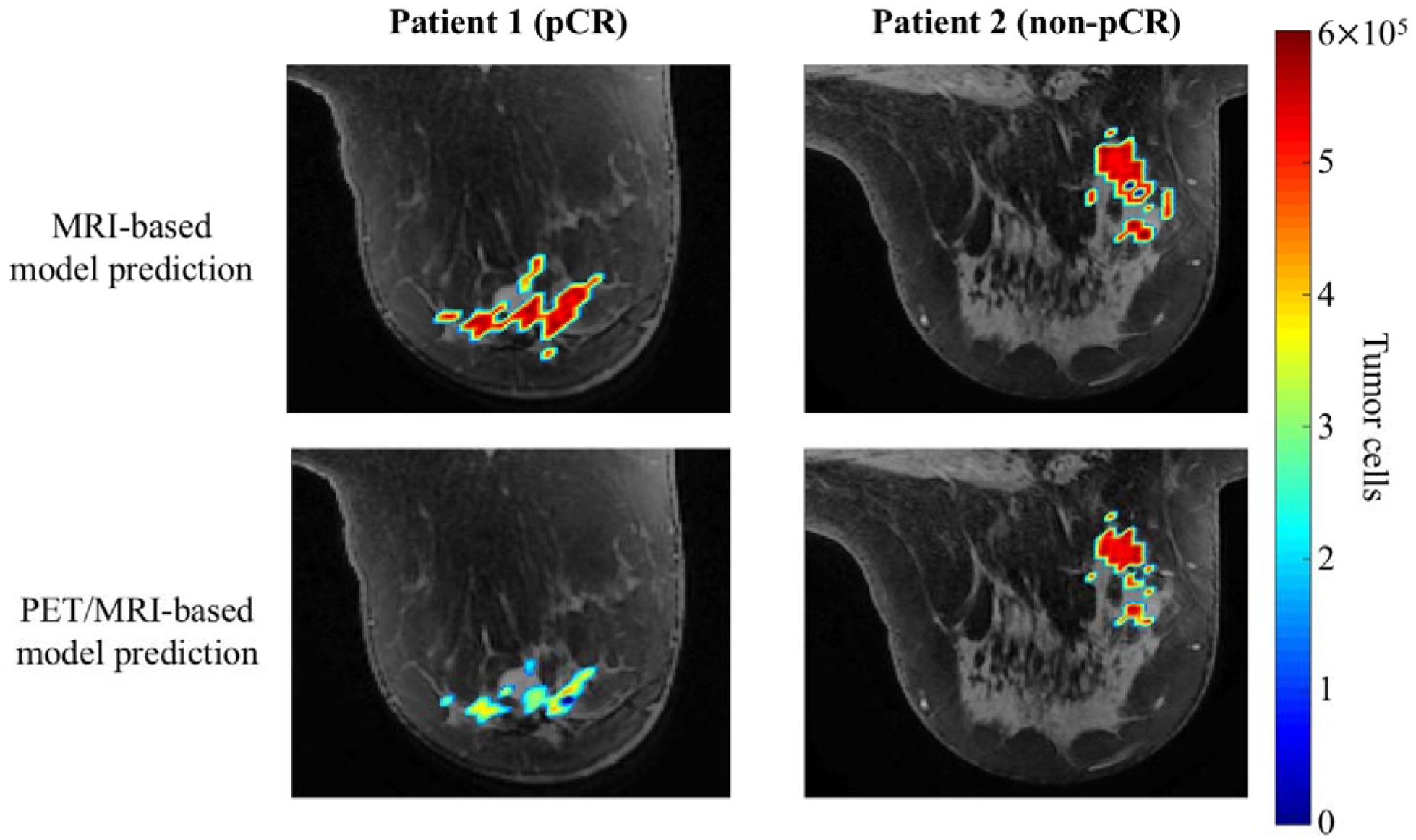
Central slice examples of the predicted tumor cellularity by the MRI-based model compared to the PET/MRI-based model. The number of tumor cells is overlaid (color) on an anatomical image of the breast (grey). At surgery patient 1’s (left column) tumor response was designated as pCR and for patient 2 (right column) the tumor response was designated as non-pCR. Notice that the PET/MRI-based model predicts overall smaller tumors and for patient 1 specifically, lower overall cellularity.

For both patients, the MRI-based model (i.e., Eqs. (1) – (4) that do not explicitly incorporate the HER2-targeted therapies) predicted that from baseline to surgery the tumors would shrink by > 50% in cellularity and volume as listed in Table 2. Specifically, the predicted overall tumor reduction by the MRI-based model was −59% and −51% for total cellularity and - 63% and −62% for volume, for patients 1 and 2, respectively. However, the model predicted that both patients will experience an increase in cellularity (203% and 86% for patients 1 and 2, respectively) and volume (46% and 37% for patients 1 and 2, respectively) from scan 2 to the time of surgery. Recall that patient 1 is the pCR patient, and patient 2 is the non-pCR patient. Thus, given the available data, the MRI-based model may prove difficult to use to distinguish between response types.

**Table 2.** Modeling results for the predicted percent change in tumor response from baseline and scan 2 to the time of surgery

| Patient → | Cellularity |  |  |  | Volume |  |  |  |
| --- | --- | --- | --- | --- | --- | --- | --- | --- |
|  | From baseline |  | From scan 2 |  | From baseline |  | From scan 2 |  |
|  | 1 | 2 | 1 | 2 | 1 | 2 | 1 | 2 |
| MRI-based model | -59% | -51% | 203% | 86% | -63% | -62% | 46% | 37% |
| PET/MRI-based model | -90% | -61% | -24% | 47% | -88% | -69% | -38% | 12% |

The expanded PET/MRI-based model also predicted for both patients that the tumors would shrink from baseline to surgery by > 60% in cellularity and volume. Specifically, the predicted overall tumor reduction by the PET/MRI-based model was −90% and −61% for total cellularity and −88% and −69% for volume, for patients 1 and 2, respectively. Note that patient 1’s tumor was predicted to have a greater decrease in total cellularity and volume when compared to the predicted response of patient 2 (by almost 30% and 20%, respectively). For the percent change from MRI 2 to the time of surgery, the model predicted opposite responses between the two patients; the model predicted the tumor cellularity and volume would decrease for the pCR patient (patient 1), while the tumor cellularity and volume would increase for the non-pCR patient (patient 2). The predicted percent change from scan 2 to the time of surgery for the PET/MRI-based model was −24% and 47% for total cellularity and −38% and 12% for volume, for patients 1 and 2, respectively. See Figure 6 for the predicted tumor response curves for total cellularity from scan 2 to the time of surgery. Notice that the MRI-based model predicted overall tumor control for both patients during therapy (oscillating between approximately 10-30% and 10-50% of the baseline total tumor cellularity for patients 1 and 2, respectively). On the other hand, the PET/MRI-based model predicted overall tumor reduction with therapy for patient 1 (progressively decreasing with each dose) and overall tumor control for patient 2 (oscillating between 20-30% percent of the baseline total tumor cellularity).

**Figure 6.**
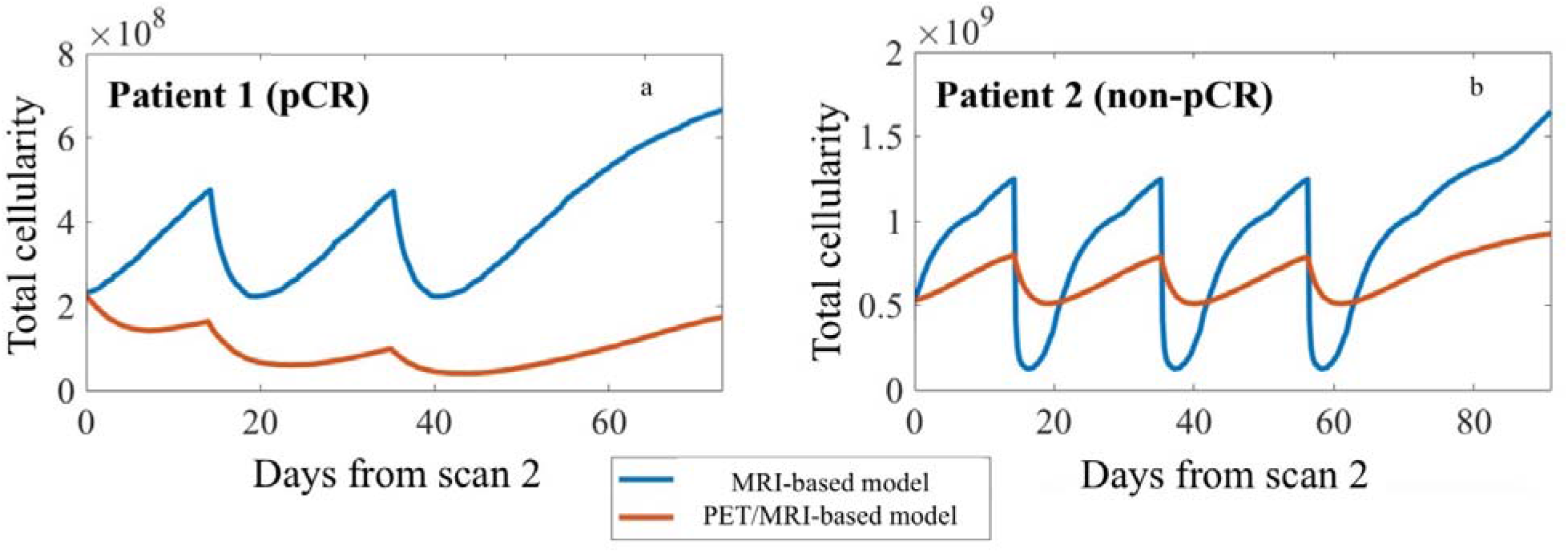
Tumor response curves for total cellularity predicted by the MRI-based (blue curve) and PET/MRI-based model (orange curve) for each patient. Each panel depicts the predictions for total tumor cellularity from the time of scan 2 (day 0) to the time of surgery (end of simulation) for patient 1 (panel a) and patient 2 (panel b). Note that patient 1 received two doses of therapy between her second scan and surgery, while patient 2 received three doses of therapy during that time. The period of tumor regrowth after the completion of NAT occurs during the time patients were no longer receiving systemic therapy. Patients 1 and 2 underwent surgery 38 and 36 days, respectively, after their last cycle of therapy. While both models predicted an oscillatory behavior in relation to when therapy was delivered, the MRI-based model (blue curves) predicted greater tumor regrowth during the refractory periods than the PET/MRI based model (orange curves). We conjecture this reflects the modified proliferation due to targeted therapy incorporated into the model.

## 4 Discussion

While there is a developing literature on integrating imaging data (and, in particular, MRI data) to generate patient-specific predictions of tumor response [16-20, 43, 53], relatively few modeling frameworks have been proposed to integrate multiple imaging modalities [54]. Despite our previous successes with predicting the response of locally-advanced breast cancer to broad-spectrum chemotherapies [20, 21], we have struggled to capture the effect of targeted therapies. As the dynamics of targeted therapy distribution are not necessarily captured by commonly available MRI techniques, we utilized PET imaging with a novel imaging agent (^64^Cu-DT) to approximate the distribution of the anti-HER2 antibody. We then compared the predictions of the MRI-based model to the PET/MRI-based model that includes the effect of targeted therapy on the proliferation of tumor cells using the PET imaging data to indicate targeted drug distribution. We found that the expanded, PET/MRI-based model gave distinctly different predictions for the two patients that more closely agreed with clinical outcome than the MRI-based model. Specifically, the MRI-based model predicted that both tumors would regrow (as indicated by both cellularity and volume) after scan 2. As surgical pathology determined that these two patients had very different outcomes (pCR and non-pCR for patient 1 and 2, respectively), the MRI-based mathematical model failed to capture key patient-specific characteristics when making its forecast. Conversely, with the incorporation of the targeted therapy data and corresponding model modification, the PET/MRI-based model provides distinct predictions for the two patients that more closely agree with clinical outcome.

To the best of our knowledge this represents the first study to combine MRI and PET data for breast cancer in a predictive mathematical model. However, previous theoretical work has shown that the linking of several different modalities of data to quantify tumor size, proliferation, metabolism, and vascularity using simple tumor growth models is not only plausible but also fundamental to capturing and understanding the biological phenomena that is cancer [55]. One example is the more recent work where MRI and PET data were combined in a clinical model of glioblastoma [54]. By using ^18^F-fluoromisonidazole (FMISO-) PET data to quantify the hypoxic areas of the tumor, the authors were able to couple this information to the effectiveness of radiation treatment and found that it decreased the error between the model’s predictions and the patient’s actual response by an order of magnitude. This result is very similar to what we observed in this breast cancer study by including the effects of targeted therapy explicitly using ^64^Cu-DT-PET.

This study has several technical limitations related to both data acquisition and mathematical modeling that should be systematically investigated in future efforts. Considering data acquisition, we are limited by the paucity of time points at which the PET and MRI are collected, and this places substantial demands on our model calibration. While additional time points prior to and during therapy would enable a more precise determination of model parameters, potentially yielding more accurate predictions, the burden placed on patients as well as the associated expense involved in additional scans, fundamentally limits the practical feasibility of acquiring additional time points. As an initial effort to incorporate additional imaging data and determine if this stratification of the modeling framework can help distinguish predictions for pCR and non-pCR patients, this small data set provides sufficient evidence to continue exploring this avenue.

Another consideration regarding the PET data, concerns the debate over which radiolabeled version of trastuzumab is most appropriate for clinical application, ^64^Cu or ^89^Zr labelled trastuzumab. The longer half-life of ^89^Zr (78 hours) makes it more appealing from an imaging perspective, but this does expose patients to roughly 2.5× the radiation dose associated with (for example) a standard ^18^F-FDG-PET scan [56, 57]. While ^64^Cu has a shorter half-life (12.7 hours), it is still sufficiently long for imaging up to 48 hours. Therefore, the copper isotope is attractive for PET imaging as well as patient safety [30, 58].

On the MRI side, a limitation we have previously discussed in detail is estimating cellularity with the *ADC* [20, 41, 59]. While there is good data indicating that the *ADC* is linearly correlated with cellularity [60], future efforts are needed to eliminate some of the ambiguity in the interpretation of *ADC*. Many other factors (cell membrane permeability [61], cell size, and tissue tortuosity [62]) in addition to cellularity can also effect the *ADC*, and therefore, estimating cellularity with *ADC* is simply an approximation.

We now discuss limitations from a modeling perspective. As it is well known that breast tumors are quite heterogeneous at all spatial and temporal scales, a limitation of the modeling approach is the assumption that there is only one tumor cell phenotype. While the spatially defined proliferation map allows the model to better capture local behavior of the tumor cells, a simplifying assumption was made that all tumor cells respond similarly to drug therapy with the global *α* and *µ* parameters. The immunohistochemistry data from the biopsy samples indicate that all of the cancerous cells (highly) overexpress HER2. Therefore, our simplifying assumption of the tumor being composed of one HER2+ cell population is supported by the fact that the biopsy data suggests that the tumors have low intratumoral variability of HER2 expression. This is also in agreement with other studies evaluating the homogeneity of HER2 staining in cancerous cells of HER2+ tumors [63, 64]. However, in patients that exhibit HER2 expression heterogeneity on the biopsy specimens, the model could be extended to include a fraction of HER2 negative cells within the tumor. The spatial distribution of HER2 negative cells in the tumor mass may be estimated with other imaging modalities, for example ^18^F-FDG, or with simplifying assumptions of well mixed populations.

Other modeling limitations stem from the incorporation of therapies into the model. First, it is important to note that assigning the delivery of drug in the mathematical model *via* CE-MRI data is only a first order approximation, not unlike other efforts that have attempted to estimate heterogeneous drug delivery to tissue [65-68]. However, we do not account for differences in the mechanisms of transport of the chemotherapy and antibodies separately or account for potential drug synergy. We also assume that the antibody concentration in the tumor is constant throughout treatment. It is well known that antibodies circulate in the body for long periods of time, evidenced by up to 7-day post administration imaging of ^89^Zr labeled trastuzumab studies [30, 69, 70]. Future studies should consider decay in antibody concentration and/or increased drug resistance. Finally, a major limitation in the model for the drug delivery in general is a non-evolving vasculature. Adding an evolving vasculature would require a significant expansion to the model (with a new governing equation for the vasculature component alone), and we have had some success with this in the pre-clinical setting but this approach requires more data than available to calibrate the model [59, 71].

In spite of these limitations, it is important to note that the model is both spatially and temporally resolved and can be calibrated almost entirely with data obtained from individual patients in the clinical setting to simulate tumor response to therapy. The ability to predict individual patient response using non-invasive imaging measures is difficult to overstate in oncology. By building clinical-mathematical frameworks capable of being calibrated and constrained with patient-specific data, we can begin to not only simulate differing patient responses but begin to identify negative outcomes early in the course of therapy. To be of any relevance, various types of therapies must be incorporated into such a mathematical system, and therefore, multiple modalities of data must be considered despite the challenges of interlacing these data types.

## 5 Conclusions

We have provided a proof-of-concept study to demonstrate how two imaging modalities (MRI and PET) can be combined in a mathematical model to provide patient-specific predictions of response to neoadjuvant chemotherapy and targeted therapy. Considering an initial two patients, we find incorporating both MRI and PET imaging data into a mathematical model allows the model to better distinguish between pCR and non-pCR outcomes. These results represent a first step to combining multiple modalities of clinically-relevant imaging data in a mathematical model for individualized predictions of therapy response.

## 6 Data Availability

The datasets generated during and/or analyzed during the current study are available from the corresponding authors on reasonable request.

## 8 Acknowledgements

We offer a sincere thank you to all the women who volunteered to participate in our studies; your strength and courage are examples for all of us. Thank you to the Baum Family and the Pacific Northwest Food Industries Circle. We thank the National Cancer Institute for support under award numbers U01CA174706, U24CA226110, U01CA142565, R01CA186193, P30CA033572; the Cancer Prevention Research Institute of Texas for funding through CPRIT RR160005, and the American Association of Physicists in Medicine for funding through the 2018 Research Seed Grant. The content is solely the responsibility of the authors and does not necessarily represent the official views of the National Institutes of Health. T.E.Y. is a CPRIT Scholar in Cancer Research.

## 9 Author Contributions

J.M., R.C.R., and T.E.Y. conceived the study. V.A., P.S., and D.A. collected the data. Authors L.T., D.S. and J.M provided expert interpretation of clinical results. A.M.J. and D.A.H processed the data. A.M.J., D.A.H., R.C.R., and T.E.Y. developed the mathematical models, and A.M.J. and D.A.H. developed the numerical methods to fit the proposed models. A.M.J. generated all the results. All authors reviewed the manuscript.

## 10 Additional Information

The authors have no competing interests.

## References

1. Chen, X., R.M. Summers, and J. Yao, Kidney tumor growth prediction by coupling reaction-diffusion and biomechanical model. IEEE Trans Biomed Eng, 2013. 60(1): p. 169–73.

2. Yuan, J. and L. Liu, Brain glioma growth model using reaction-diffusion equation with viscous stress tensor on brain MR images. Magnetic Resonance Imaging, 2016. 34 (2): p. 114–9.

3. Rockne, R., et al., Predicting the efficacy of radiotherapy in individual glioblastoma patients in vivo: a mathematical modeling approach. Phys Med Biol, 2010. 55(12): p. 3271–85.

4. Clatz, O., et al., Realistic simulation of the 3-D growth of brain tumors in MR images coupling diffusion with biomechanical deformation. Ieee Transactions on Medical Imaging, 2005. 24(10): p. 1334–1346.

5. Baldock, A.L., et al., From patient-specific mathematical neuro-oncology to precision medicine. Front Oncol, 2013. 3: p. 62.

6. Mi, H., et al., Prediction of lung tumor evolution during radiotherapy in individual patients with PET. IEEE Trans Med Imaging, 2014. 33(4): p. 995–1003.

7. Mi, H., et al., Joint tumor growth prediction and tumor segmentation on therapeutic follow-up PET images. Med Image Anal, 2015. 23(1): p. 84–91.

8. Liu, Y.X., et al., Patient specific tumor growth prediction using multimodal images. Medical Image Analysis, 2014. 18(3): p. 555–566.

9. Wong, K.C., et al., Pancreatic Tumor Growth Prediction with Multiplicative Growth and Image-Derived Motion. Inf Process Med Imaging, 2015. 24: p. 501–13.

10. Liu, Y., et al., Multimodal image driven patient specific tumor growth modeling. Med Image Comput Comput Assist Interv, 2013. 16(Pt 3): p. 283–90.

11. Wong, K.C., et al., Tumor growth prediction with reaction-diffusion and hyperelastic biomechanical model by physiological data fusion. Med Image Anal, 2015. 25(1): p. 72–85.

12. Hormuth, D.A., et al., Translating preclinical MRI methods to clinical oncology. J Magn Reson Imaging, 2019.

13. ME, P., PET: Molecular Imaging and its Biological Applications. 2004: Springer.

14. Atuegwu, N.C., et al., Integration of diffusion-weighted MRI data and a simple mathematical model to predict breast tumor cellularity during neoadjuvant chemotherapy. Magn Reson Med, 2011. 66(6): p. 1689–96.

15. Atuegwu, N.C., et al., Incorporation of diffusion-weighted magnetic resonance imaging data into a simple mathematical model of tumor growth. Physics in Medicine and Biology, 2012. 57(1): p. 225–240.

16. Atuegwu, N.C., et al., Parameterizing the Logistic Model of Tumor Growth by DW-MRI and DCE-MRI Data to Predict Treatment Response and Changes in Breast Cancer Cellularity during Neoadjuvant Chemotherapy. Translational Oncology, 2013. 6(3): p. 256–264.

17. Weis, J.A., et al., A mechanically coupled reaction-diffusion model for predicting the response of breast tumors to neoadjuvant chemotherapy. Physics in Medicine and Biology, 2013. 58(17): p. 5851–5866.

18. Weis, J.A., et al., Predicting the Response of Breast Cancer to Neoadjuvant Therapy Using a Mechanically Coupled Reaction-Diffusion Model. Cancer Research, 2015. 75(22): p. 4697–4707.

19. Weis, J.A., M.I. Miga, and T.E. Yankeelov, Three-dimensional image-based mechanical modeling for predicting the response of breast cancer to neoadjuvant therapy. Computer Methods in Applied Mechanics and Engineering, 2017. 314: p. 494–512.

20. Jarrett, A.M., et al., Incorporating drug delivery into an imaging-driven, mechanics-coupled reaction diffusion model for predicting the response of breast cancer to neoadjuvant chemotherapy: theory and preliminary clinical results. Phys Med Biol, 2018. 63(10): p. 105015.

21. Jarrett, A., et al., Optimizing neoadjuvant regimens for individual breast cancer patients generated by a mathematical model utilizing quantitative magnetic resonance imaging data: Preliminary results. Cancer Research, 2020.

22. Gonzalez-Angulo, A.M., et al., High risk of recurrence for patients with breast cancer who have human epidermal growth factor receptor 2-positive, node-negative tumors 1 cm or smaller. J Clin Oncol, 2009. 27(34): p. 5700–6.

23. Rosen, L.S., H.L. Ashurst, and L. Chap, Targeting signal transduction pathways in metastatic breast cancer: a comprehensive review. Oncologist, 2010. 15(3): p. 216–35.

24. Henry, K.E., G.A. Ulaner, and J.S. Lewis, Clinical Potential of Human Epidermal Growth Factor Receptor 2 and Human Epidermal Growth Factor Receptor 3 Imaging in Breast Cancer. PET Clin, 2018. 13(3): p. 423–435.

25. Williams, J.M., et al., Comparison of prone versus supine 18F-FDG-PET of locally advanced breast cancer: Phantom and preliminary clinical studies. Med Phys, 2015. 42(7): p. 3801–13.

26. Abramson, R.G., et al., Prone Versus Supine Breast FDG-PET/CT for Assessing Locoregional Disease Distribution in Locally Advanced Breast Cancer. Acad Radiol, 2015. 22(7): p. 853–9.

27. Atuegwu, N.C., et al., Longitudinal, intermodality registration of quantitative breast PET and MRI data acquired before and during neoadjuvant chemotherapy: preliminary results. Med Phys, 2014. 41(5): p. 052302.

28. Li, X., et al., An algorithm for longitudinal registration of PET/CT images acquired during neoadjuvant chemotherapy in breast cancer: preliminary results. EJNMMI Res, 2012. 2(1): p. 62.

29. Mortimer, J.E., et al., Tumor Uptake of ^64^Cu-DOTA-Trastuzumab in Patients with Metastatic Breast Cancer. J Nucl Med, 2018. 59(1): p. 38–43.

30. Mortimer, J.E., et al., Functional imaging of human epidermal growth factor receptor 2-positive metastatic breast cancer using (64)Cu-DOTA-trastuzumab PET. J Nucl Med, 2014. 55(1): p. 23–9.

31. Whisenant, J.G., et al., Assessing reproducibility of diffusion-weighted magnetic resonance imaging studies in a murine model of HER2+ breast cancer. Magn Reson Imaging, 2014. 32(3): p. 245–9.

32. Wu, C., et al., Quantitative analysis of vascular properties derived from ultrafast DCE-MRI to discriminate malignant and benign breast tumors. Magn Reson Med, 2018.

33. Li, X., et al., Validation of an algorithm for the nonrigid registration of longitudinal breast MR images using realistic phantoms. Med Phys, 2010. 37(6): p. 2541–52.

34. Klein, S., et al., elastix: a toolbox for intensity-based medical image registration. IEEE Trans Med Imaging, 2010. 29(1): p. 196–205.

35. Shamonin, D.P., et al., Fast parallel image registration on CPU and GPU for diagnostic classification of Alzheimer’s disease. Front Neuroinform, 2013. 7: p. 50.

36. Hagmann, P., et al., Understanding diffusion MR imaging techniques: from scalar diffusion-weighted imaging to diffusion tensor imaging and beyond. Radiographics, 2006. 26 Suppl 1: p. S205–23.

37. Anderson, A.W., et al., Effects of cell volume fraction changes on apparent diffusion in human cells. Magn Reson Imaging, 2000. 18(6): p. 689–95.

38. Martin, I., et al., Computer-based technique for cell aggregation analysis and cell aggregation in in vitro chondrogenesis. Cytometry, 1997. 28(2): p. 141–6.

39. McKnight, A.L., et al., MR elastography of breast cancer: preliminary results. AJR Am J Roentgenol, 2002. 178(6): p. 1411–7.

40. Hormuth, D.A., et al., Predicting in vivo glioma growth with the reaction diffusion equation constrained by quantitative magnetic resonance imaging data. Physical Biology, 2015. 12(4).

41. Hormuth, D.A., et al., A mechanically coupled reaction-diffusion model that incorporates intra-tumoural heterogeneity to predict in vivo glioma growth. J R Soc Interface, 2017. 14(128).

42. Hormuth, D.A., et al., Mechanically Coupled Reaction-Diffusion Model to Predict Glioma Growth: Methodological Details. Methods Mol Biol, 2018. 1711: p. 225–241.

43. Hormuth, D.A., et al., Mechanism-based Modeling of Tumor Growth and Treatment Response Constrained by Multiparametric Imaging Data. Jco Clinical Cancer Informatics, 2019(3): p. 10.

44. Hormuth II, D., et al., Predicting in vivo tumor growth and angiogenesis with an MRI calibrated biophysical model Neuro-Oncology, 2017. 19(suppl_6): p. vi23.

45. Hormuth II DA, et al., Biophysical modeling of in vivo glioma response following whole brain radiotherapy in a murine model of brain cancer. International Journal of Radiation Oncology • Biology • Physics 2018.

46. Barpe, D.R., D.D. Rosa, and P.E. Froehlich, Pharmacokinetic evaluation of doxorubicin plasma levels in normal and overweight patients with breast cancer and simulation of dose adjustment by different indexes of body mass. Eur J Pharm Sci, 2010. 41(3-4): p. 458–63.

47. van der Vijgh, W.J., Clinical pharmacokinetics of carboplatin. Clin Pharmacokinet, 1991. 21(4): p. 242–61.

48. Mori, T., et al., Retention of paclitaxel in cancer cells for 1 week in vivo and in vitro. Cancer Chemother Pharmacol, 2006. 58(5): p. 665–72.

49. Tew, K., Paclitaxel. Reference Module in Biomedical Sciences, 2016.

50. Yang, L., et al., Pharmacokinetics and safety of cyclophosphamide and docetaxel in a hemodialysis patient with early stage breast cancer: a case report. BMC Cancer, 2015. 15: p. 917.

51. Powis, G., et al., Effect of body weight on the pharmacokinetics of cyclophosphamide in breast cancer patients. Cancer Chemother Pharmacol, 1987. 20(3): p. 219–22.

52. Jarrett, A.M., et al., Experimentally-driven mathematical modeling to improve combination targeted and cytotoxic therapy for HER2+ breast cancer. Sci Rep, 2019. 9(1): p. 12830.

53. Jarrett, A.M., et al., Abstract P2-16-17: Optimizing neoadjuvant regimens for individual breast cancer patients generated by a mathematical model utilizing quantitative magnetic resonance imaging data: Preliminary results, in SABCS. 2019: San Antonio, TX, USA.

54. Rockne, R.C., et al., A patient-specific computational model of hypoxia-modulated radiation resistance in glioblastoma using 18F-FMISO-PET. J R Soc Interface, 2015. 12(103).

55. Atuegwu, N.C., J.C. Gore, and T.E. Yankeelov, The integration of quantitative multi-modality imaging data into mathematical models of tumors. Phys Med Biol, 2010. 55(9): p. 2429–49.

56. Tamura, K., et al., 64Cu-DOTA-trastuzumab PET imaging in patients with HER2-positive breast cancer. J Nucl Med, 2013. 54(11): p. 1869–75.

57. Schjoeth-Eskesen, C., et al., [(64) Cu]-labelled trastuzumab: optimisation of labelling by DOTA and NODAGA conjugation and initial evaluation in mice. J Labelled Comp Radiopharm, 2015. 58(6): p. 227–33.

58. Caserta, E., et al., Copper 64-labeled daratumumab as a PET/CT imaging tracer for multiple myeloma. Blood, 2018. 131(7): p. 741–745.

59. Hormuth, D.A., et al., Calibrating a Predictive Model of Tumor Growth and Angiogenesis with Quantitative MRI. Ann Biomed Eng, 2019. 47(7): p. 1539–1551.

60. Barnes, S.L., et al., Correlation of tumor characteristics derived from DCE-MRI and DW-MRI with histology in murine models of breast cancer. NMR Biomed, 2015. 28(10): p. 1345–56.

61. Latour, L.L., et al., Time-dependent diffusion of water in a biological model system. Proc Natl Acad Sci U S A, 1994. 91(4): p. 1229–33.

62. van der Toorn, A., et al., Dynamic changes in water ADC, energy metabolism, extracellular space volume, and tortuosity in neonatal rat brain during global ischemia. Magn Reson Med, 1996. 36(1): p. 52–60.

63. Press, M.F., et al., Her-2/neu expression in node-negative breast cancer: direct tissue quantitation by computerized image analysis and association of overexpression with increased risk of recurrent disease. Cancer Res, 1993. 53(20): p. 4960–70.

64. Press, M.F., et al., HER-2 gene amplification, HER-2 and epidermal growth factor receptor mRNA and protein expression, and lapatinib efficacy in women with metastatic breast cancer. Clin Cancer Res, 2008. 14(23): p. 7861–70.

65. Swanson, K.R., E.C. Alvord, and J.D. Murray, Quantifying efficacy of chemotherapy of brain tumors with homogeneous and heterogeneous drug delivery. Acta Biotheoretica, 2002. 50(4): p. 223–237.

66. Kim, M., R.J. Gillies, and K.A. Rejniak, Current advances in mathematical modeling of anti-cancer drug penetration into tumor tissues. Front Oncol, 2013. 3: p. 278.

67. Owen, M.R., et al., Mathematical modeling predicts synergistic antitumor effects of combining a macrophage-based, hypoxia-targeted gene therapy with chemotherapy. Cancer Res, 2011. 71(8): p. 2826–37.

68. Shah, A.B., K.A. Rejniak, and J.L. Gevertz, Limiting the Development of Anti-cancer Drug Resistance in a Spatial Model of Micrometastases. Mathematical Biosciences and Engineering, 2016. 13(6): p. 1185–1206.

69. Laforest, R., et al., [89 Zr]Trastuzumab: Evaluation of Radiation Dosimetry, Safety and Optimal Imaging Parameters in Women with HER2-Positive Breast Cancer. Mol Imaging Biol, 2016. 18(6): p. 952–959.

70. O’Donoghue, J.A., et al., Pharmacokinetics, biodistribution, and radiation dosimetry for ^89^Zr-trastuzumab in patients with esophagogastric cancer. J Nucl Med, 2018. 59(1): p. 161–166.

71. Hormuth, D.A., A.M. Jarrett, and T.E. Yankeelov, Forecasting tumor and vasculature response dynamics to radiation therapy via image based mathematical modeling. Radiat Oncol, 2020. 15(1): p. 4.

